# Associations Among Changes in Inflammatory Biomarkers, Pain Intensity, and Health-Related Quality of Life Following a 12-Week Aerobic Exercise Programme in Individuals with Non-Specific Chronic Low Back Pain

**DOI:** 10.64898/2026.06.21.26356167

**Authors:** Nweke Vincent Chinonso, Kadiree Ekundayo Fatai, Anelechi Kenneth Madume, Chidiebele P Ojukwu, Adaeze Imelda Onyekwelu, Akachukwu O. Nwosu, Nweke Queeneth kadilobari, Nweke Augustine Chidera, Charles I. Ezema

## Abstract

**Background:** Non-specific chronic low back pain (NSCLBP) is associated with persistent pain, reduced health-related quality of life (HRQoL), and low-grade systemic inflammation. This study examined associations among changes in inflammatory biomarkers, pain intensity, and HRQoL following a 12-week aerobic exercise programme.

**Methods:** This secondary analysis used data from a randomized controlled trial involving 41 participants with NSCLBP (intervention, n = 21; control, n = 20). Participants received either supervised aerobic exercise plus health education or health education alone for 12 weeks. Change scores for tumour necrosis factor-alpha (TNF-α), interleukin-6 (IL-6), high-sensitivity C-reactive protein (hs-CRP), pain intensity, and HRQoL domains were analysed using correlation and multiple regression analyses.

**Results:** Improvements in IL-6 (r = 0.434, p = 0.005) and hs-CRP (r = 0.444, p = 0.004) were significantly associated with improvements in pain intensity. No significant associations were observed between biomarker changes and HRQoL domains. Treatment allocation was the strongest independent predictor of improvement in physical HRQoL (β = 0.492, p = 0.017) and pain intensity (β = −0.512, p = 0.006).

**Conclusions:** Improvements in IL-6 and hs-CRP were associated with reductions in pain intensity but not with improvements in HRQoL. Treatment allocation was the strongest predictor of clinical improvement, suggesting that mechanisms beyond systemic inflammation may contribute to the benefits of aerobic exercise in NSCLBP.

**Plain Language Summary:** Non-specific chronic low back pain (NSCLBP) is a common condition that can cause persistent pain, reduce daily functioning, and negatively affect quality of life. Research suggests that low-grade inflammation may contribute to pain persistence, but it remains unclear how changes in inflammation relate to improvements in pain and health-related quality of life (HRQoL) following exercise-based rehabilitation.

This study examined the relationships among changes in inflammatory biomarkers, pain intensity, and HRQoL following a 12-week aerobic exercise programme in people with NSCLBP. Forty-one participants completed either a supervised aerobic exercise programme combined with health education or health education alone. Changes in inflammatory biomarkers (IL-6, hs-CRP, and TNF-α), pain intensity, and HRQoL were assessed over the intervention period.

The findings showed that improvements in IL-6 and hs-CRP were associated with reductions in pain intensity. However, changes in inflammatory biomarkers were not significantly associated with improvements in HRQoL. The strongest predictor of improvements in both pain and physical HRQoL was participation in the aerobic exercise programme.

These findings suggest that aerobic exercise can improve outcomes in people with NSCLBP, but the benefits may not be explained solely by reductions in inflammation. Other factors, including physical, psychological, and behavioural changes associated with exercise, may also contribute to recovery and improved well-being.

## Background

Low back pain (LBP) is among the most prevalent musculoskeletal disorders worldwide and remains the leading cause of years lived with disability (YLDs) [1]. Approximately 85–90% of LBP cases are classified as non-specific low back pain, indicating that symptoms cannot be attributed to a specific pathological condition such as fracture, infection, malignancy, or inflammatory disease [2,3]. Chronic low back pain (CLBP), defined as pain persisting for more than 12 weeks, contributes substantially to disability, healthcare utilisation, productivity loss, and impaired quality of life [2]. The burden of CLBP continues to rise globally, particularly in low-and middle-income countries such as Nigeria, owing to population growth, urbanisation, ageing, and increasingly sedentary lifestyles [1].

The aetiology and persistence of non-specific chronic low back pain (NSCLBP) are multifactorial and are best understood through the biopsychosocial model, which recognises the interaction of biological, psychological, and social factors in shaping pain experiences and disability [6]. Among the biological mechanisms implicated in chronic pain, systemic inflammation has received increasing attention because of its role in pain sensitisation and symptom persistence [4,5]. Elevated concentrations of pro-inflammatory biomarkers, including tumour necrosis factor-alpha (TNF-α), interleukin-6 (IL-6), and C-reactive protein (CRP), have been reported among individuals with NSCLBP and have been associated with greater pain severity, disability, and poorer clinical outcomes [4,5].

Inflammatory mediators contribute to chronic pain through multiple mechanisms, including peripheral nociceptor sensitisation, neuroimmune interactions, and dysregulation of central pain-processing pathways [4,5]. Persistent low-grade inflammation may therefore sustain pain beyond the initial tissue insult and facilitate the transition from acute to chronic pain states [4]. Emerging evidence further suggests that systemic inflammation may adversely influence broader health outcomes through its effects on pain perception, physical functioning, and symptom burden [9,10]. Consequently, individuals with higher inflammatory activity may experience greater pain intensity and more pronounced functional limitations.

Pain intensity is a central determinant of health outcomes in NSCLBP because it directly affects physical functioning, participation in daily activities, psychological well-being, and social engagement [2]. Persistent pain is associated with fear-avoidance behaviours, reduced physical activity, emotional distress, anxiety, depression, and social withdrawal, all of which contribute to poorer health-related quality of life [6,7]. Individuals with chronic low back pain consistently report lower quality-of-life scores across physical, psychological, social, and environmental domains than healthy populations [7,8].

Aerobic exercise is widely recommended as a first-line intervention for NSCLBP because of its beneficial effects on pain, physical function, and overall health status. In addition to improving physical conditioning, aerobic exercise may modulate inflammatory processes through reductions in systemic inflammatory activity and improvements in physiological function. Consequently, changes in inflammatory biomarkers may occur alongside changes in pain intensity and quality of life following exercise-based rehabilitation.

Although systemic inflammation, pain intensity, and quality of life have each been independently associated with NSCLBP, relatively few studies have examined how changes in these outcomes are related following rehabilitation interventions. Understanding the associations among changes in inflammatory biomarkers, pain intensity, and quality of life may provide insight into the biological and clinical responses accompanying exercise-based rehabilitation in individuals with chronic low back pain.

Evidence regarding these interrelationships remains limited, particularly within Sub-Saharan African populations where environmental, occupational, and healthcare contexts may differ from those reported in high-income countries [4,5]. Improved understanding of these associations may help identify clinically relevant targets for rehabilitation interventions aimed at reducing symptom burden and improving overall well-being in individuals with NSCLBP.

Therefore, this study aimed to examine the associations among changes in systemic inflammatory biomarkers, pain intensity, and health-related quality of life following a 12-week aerobic exercise programme in individuals with NSCLBP. Specifically, the study investigated the relationships among changes in serum concentrations of TNF-α, IL-6, and high-sensitivity C-reactive protein (hs-CRP), pain intensity, and quality-of-life outcomes. It was hypothesised that improvements in inflammatory biomarkers would be associated with improvements in pain intensity and health-related quality of life following the intervention period.

## Methods

### Study Design

This study examined the associations among changes in systemic inflammatory biomarkers, pain intensity, and quality of life following a 12-week aerobic exercise programme in individuals with non-specific chronic low back pain (NSCLBP). Data were derived from a parallel-group randomized controlled trial that evaluated the effects of aerobic exercise on inflammatory biomarkers, pain intensity, and health-related quality of life. Participants were randomly assigned to either an aerobic exercise plus health education group or a health education-only control group. Outcome assessments were conducted at baseline (Week 0), Week 8, and Week 12.

The present analysis focused on changes in serum concentrations of tumour necrosis factor-alpha (TNF-α), interleukin-6 (IL-6), and high-sensitivity C-reactive protein (hs-CRP), pain intensity, and quality-of-life outcomes over the intervention period. Associations among these variables were examined using correlation and regression analyses. Changes in inflammatory biomarkers, pain intensity, and quality of life were calculated as the difference between baseline and Week 12 values.

The parent randomized controlled trial was conducted in accordance with the Consolidated Standards of Reporting Trials (CONSORT) 2010 Statement [11,12]. The trial was retrospectively registered with the Pan African Clinical Trials Registry (PACTR; Registration Number: PACTR202606887164888) on 11 June 2026 due to administrative delays. The study protocol, eligibility criteria, intervention procedures, outcome measures, and statistical analysis plan were finalised before participant recruitment and remained unchanged following registration.

### Study Setting

The study was conducted at the Physiotherapy Department of Rivers State University Teaching Hospital (RSUTH), Port Harcourt, Rivers State, Nigeria. RSUTH is a tertiary healthcare institution that provides specialist physiotherapy services and serves as a referral centre for patients within Rivers State and neighbouring states in the Niger Delta region. Participant recruitment, intervention delivery, and outcome assessments were conducted within the Physiotherapy Department and affiliated diagnostic laboratories.

### Sample Size Determination

The sample size for the parent randomized controlled trial was determined a priori using G*Power version 3.1 [13] for a mixed-design repeated-measures analysis of variance (ANOVA). The calculation was based on a moderate effect size (Cohen’s f = 0.25), a significance level of 5% (α = 0.05), and a statistical power of 80% (1−β = 0.80). The analysis indicated that a minimum sample size of 36 participants was required. To account for a potential attrition rate of 20%, the target sample size was increased to 44 participants. A total of 41 participants completed the study and were included in the analyses.

Because the sample size was determined to detect intervention effects in the parent randomized controlled trial rather than to examine associations among changes in inflammatory biomarkers, pain intensity, and quality-of-life outcomes, the findings of the present secondary analysis should be interpreted with appropriate caution. Consequently, the study may have been underpowered to detect weak associations among the variables examined.

### Interim Analyses and Stopping Guidelines

No interim analyses or formal stopping guidelines were planned because of the short duration and low-risk nature of the intervention.

### Participant Recruitment and Screening

Participants were recruited consecutively from the Physiotherapy Department of Rivers State University Teaching Hospital (RSUTH) through referrals from physiotherapists and physicians involved in the management of chronic low back pain. Eligibility was determined through medical history review, physical examination, and assessment against predefined eligibility criteria. Written informed consent was obtained from all participants prior to enrolment. Participant flow through the parent randomized controlled trial is presented in the CONSORT flow diagram (Figure 1).

**Figure 1:**
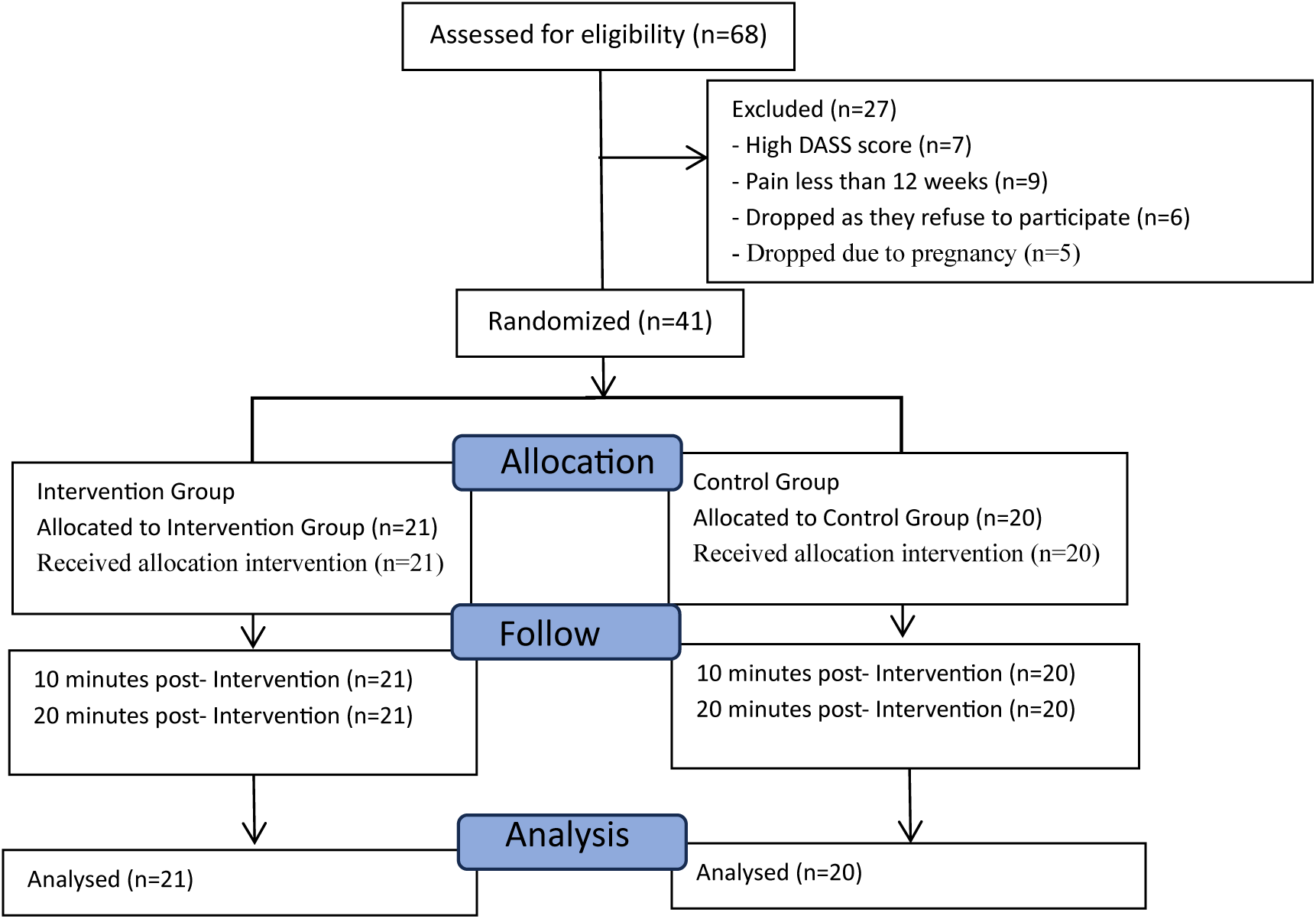
CONSORT Flow Diagram of Participant Recruitment, Randomization, Allocation, Follow-up, and Analysis.

### Eligibility Criteria

#### Inclusion Criteria

Participants were eligible for inclusion if they were aged 18 years or older; had a clinical diagnosis of non-specific chronic low back pain persisting for at least 12 weeks; were medically stable and able to participate in moderate-intensity aerobic exercise; were willing to comply with all study procedures; and provided written informed consent prior to participation.

#### Exclusion Criteria

Participants were excluded if they had a history of spinal surgery; presented with signs of lumbar radiculopathy, spinal stenosis, nerve root compression, or other specific spinal pathologies; had inflammatory rheumatic disorders, spinal infection, vertebral fracture, malignancy, or other systemic inflammatory conditions; were pregnant; demonstrated cognitive impairment that could limit participation or compliance with study procedures; or had uncontrolled cardiovascular, respiratory, neurological, or metabolic disorders that contraindicated participation in aerobic exercise.

#### Eligibility Criteria for Intervention Providers

The aerobic exercise intervention was delivered by licensed physiotherapists with clinical experience in musculoskeletal rehabilitation. All intervention providers received orientation regarding the study protocol prior to participant enrolment.

#### Equity, Diversity and Inclusion Statement

The research team considered equity, diversity, and inclusion throughout the study design and conduct. Eligibility criteria were based on clinical characteristics rather than sex, ethnicity, religion, socioeconomic status, or other demographic factors. Both male and female participants were eligible for recruitment. Participants were recruited consecutively from a routine clinical population attending a tertiary healthcare facility, and no groups were intentionally excluded from participation beyond the predefined clinical eligibility criteria.

#### Randomization and Allocation Concealment

Following baseline assessment, participants were randomly allocated to either the intervention group or the control group using a computer-generated block randomization sequence with a block size of four and an allocation ratio of 1:1. The random allocation sequence was generated by an independent researcher using a computerized random-number generator. The independent researcher was not involved in participant recruitment, enrolment, intervention delivery, outcome assessment, or data analysis.

Allocation concealment was maintained using sequentially numbered, sealed, opaque envelopes containing group assignments. The envelopes were prepared by the independent researcher and opened only after completion of baseline assessment, thereby preventing foreknowledge of treatment allocation and minimising selection bias.

#### Blinding

Due to the nature of the intervention, blinding of participants and treating physiotherapists was not feasible. However, laboratory personnel responsible for analysing blood samples and research assistants involved in outcome assessment remained blinded to group allocation throughout the study period.

### Intervention Procedures

#### Aerobic Exercise Group

Participants allocated to the intervention group completed a supervised aerobic exercise programme using a Monark Ergomedic 828E cycle ergometer (Monark Exercise AB, Vansbro, Sweden). The programme was prescribed according to the Frequency, Intensity, Time, and Type (FITT) principle and established exercise training guidelines [14,15]. Exercise sessions were conducted three times weekly for 12 weeks under the supervision of a physiotherapist. Intensity was prescribed using Heart Rate Reserve (HRR), calculated using the Karvonen formula [16], and monitored using Borg’s Rating of Perceived Exertion (RPE) Scale [17].

Each session consisted of a 5–10-minute warm-up of low-intensity cycling and flexibility exercises, followed by 30–45 minutes of aerobic cycling. Exercise intensity progressed from 40–50% HRR during Weeks 1–2 to 60–70% HRR by Week 6 and was maintained thereafter. Sessions concluded with 10–15 minutes of low-intensity cycling, stretching, and relaxation exercises. Attendance and adherence were monitored throughout the intervention period.

#### Control Group

Participants assigned to the control group received health education on the nature and management of non-specific chronic low back pain, including posture, activity modification, back care principles, and self-management strategies. Participants were instructed to maintain their usual activities and refrain from initiating structured exercise programmes during the study period. Following study completion, control participants were offered physiotherapy management.

#### Outcome Measures

Outcome assessments were conducted at baseline (Week 0), Week 8, and Week 12. The present analysis examined associations among changes in systemic inflammatory biomarkers, pain intensity, and health-related quality of life following the intervention period.

#### Systemic Inflammation

Systemic inflammation was assessed using serum concentrations of tumour necrosis factor-alpha (TNF-α), interleukin-6 (IL-6), and high-sensitivity C-reactive protein (hs-CRP). Venous blood samples were collected by trained laboratory personnel under standardised conditions at least 48 hours after the participant’s most recent exercise session to minimise the influence of acute exercise-induced inflammatory responses. Biomarker concentrations were measured using commercially available high-sensitivity enzyme-linked immunosorbent assay (ELISA) kits according to the manufacturers’ instructions. Laboratory personnel remained blinded to group allocation throughout the analytical process.

#### Pain Intensity

Pain intensity was assessed using the Visual Analogue Scale (VAS), a valid and reliable measure of pain severity in individuals with chronic pain [18]. Participants rated their average pain intensity on a 10-cm horizontal line anchored by 0 (“no pain”) and 10 (“worst imaginable pain”), with higher scores indicating greater pain intensity.

#### Quality of Life

Health-related quality of life was assessed using the World Health Organization Quality of Life Brief Questionnaire (WHOQOL-BREF) [19]. The WHOQOL-BREF evaluates four domains of quality of life, namely physical health, psychological health, social relationships, and environmental health. Domain scores were calculated according to the World Health Organization scoring guidelines, with higher scores indicating better perceived quality of life.

#### Harms Assessment

Adverse events were monitored systematically throughout the intervention period. An adverse event was defined as any undesirable medical occurrence, symptom, injury, or health-related complaint arising during study participation, irrespective of whether it was considered related to the intervention. Participants were asked to report any adverse events during exercise sessions and follow-up assessments. Physiotherapists supervising the intervention documented all reported events, including musculoskeletal discomfort, dizziness, fatigue, cardiovascular symptoms, or any other unexpected occurrence. The severity and potential relationship to the intervention were recorded and reviewed by the research team.

#### Ethical Considerations

Ethical approval for the study was obtained from the Rivers State University Teaching Hospital Health Research Ethics Committee (Approval No.: RSUTH/REC/2025897). Administrative permission to conduct the study was obtained from Rivers State University Teaching Hospital. Written informed consent was obtained from all participants prior to enrolment. The study was conducted in accordance with the ethical principles outlined in the Declaration of Helsinki.

#### Statistical Analysis

Data were analysed using IBM SPSS Statistics version 24.0 (IBM Corp., Armonk, NY, USA). All participants who completed baseline and follow-up assessments were included in the analyses. As there were no missing outcome data, no imputation procedures were required.

Descriptive statistics were computed for all study variables. Continuous variables were summarised using means and standard deviations (SD), whereas categorical variables were presented as frequencies and percentages. Data normality was assessed using the Shapiro–Wilk test, histograms, and Q–Q plots.

The present analyses were based on changes observed between baseline and Week 12 to capture the overall effect of the intervention period. Although inflammatory biomarkers and pain intensity were also assessed at Week 8, these intermediate assessments were not included in the present analyses. Change scores were calculated by subtracting baseline values from Week 12 values. Thus, changes in TNF-α, IL-6, hs-CRP, pain intensity, and WHOQOL-BREF domain scores represented improvements or deteriorations occurring over the intervention period. Change scores were selected to evaluate whether improvements in inflammatory biomarkers were associated with concurrent improvements in pain intensity and quality of life over the intervention period.

Associations among changes in inflammatory biomarkers, pain intensity, and quality-of-life outcomes were examined using Pearson’s correlation coefficients for normally distributed variables and Spearman’s rank correlation coefficients for non-normally distributed variables.

Multiple linear regression analyses were performed to identify predictors of changes in pain intensity and quality-of-life outcomes. Variables demonstrating significant or clinically relevant associations in the correlation analyses were entered into the regression models. Age and treatment allocation were included as covariates to account for potential confounding effects. Assumptions of linearity, homoscedasticity, normality of residuals, independence of errors, and multicollinearity were assessed before model interpretation. Multicollinearity was evaluated using variance inflation factors (VIFs) and tolerance statistics. VIF values ranged from 1.09 to 2.14, indicating no evidence of problematic multicollinearity among the predictor variables.

For regression analyses examining predictors of physical quality-of-life improvement, changes in inflammatory biomarkers and pain intensity were entered as independent variables, with age and treatment allocation included as covariates. A separate regression model was conducted to identify predictors of changes in pain intensity.

Regression coefficients, 95% confidence intervals, R² values, adjusted R² values, and associated p-values were reported. Statistical significance was set at p < 0.05 for all analyses.

## Results

### Participant Flow

Participant recruitment, allocation, follow-up, and analysis are presented in the CONSORT flow diagram (Figure 1). A total of 44 individuals were assessed for eligibility, of whom three were excluded prior to randomization. The remaining 41 participants met the eligibility criteria and were randomly allocated to either the aerobic exercise plus health education group (n = 21) or the health education-only control group (n = 20). No participants were lost to follow-up or withdrew during the study period. Consequently, all randomized participants completed the 12-week intervention and were included in the final analyses.

### Adverse Events

No serious adverse events, exercise-related injuries, or unintended effects were reported during the intervention period in either group.

### Intervention Adherence

Participants in the aerobic exercise group demonstrated complete adherence to the intervention protocol. All participants attended all scheduled exercise sessions throughout the 12-week intervention period, corresponding to an adherence rate of 100%.

### Participant Characteristics

Baseline demographic and clinical characteristics are presented in Table 1. A total of 41 participants were included in the study (intervention, n = 21; control, n = 20). The groups were generally comparable at baseline. However, participants in the control group were significantly older than those in the intervention group (39.55 ± 4.50 vs. 35.19 ± 7.92 years, p = 0.037, d = 0.67), and baseline IL-6 concentrations were significantly higher in the intervention group (6.96 ± 1.43 vs. 4.37 ± 1.66 pg/mL, p < 0.001, d = 1.67). No other significant between-group differences were observed for duration of symptoms, anthropometric characteristics, pain intensity, hs-CRP, TNF-α, or quality-of-life domain scores (all p > 0.05).

**Table 1.**
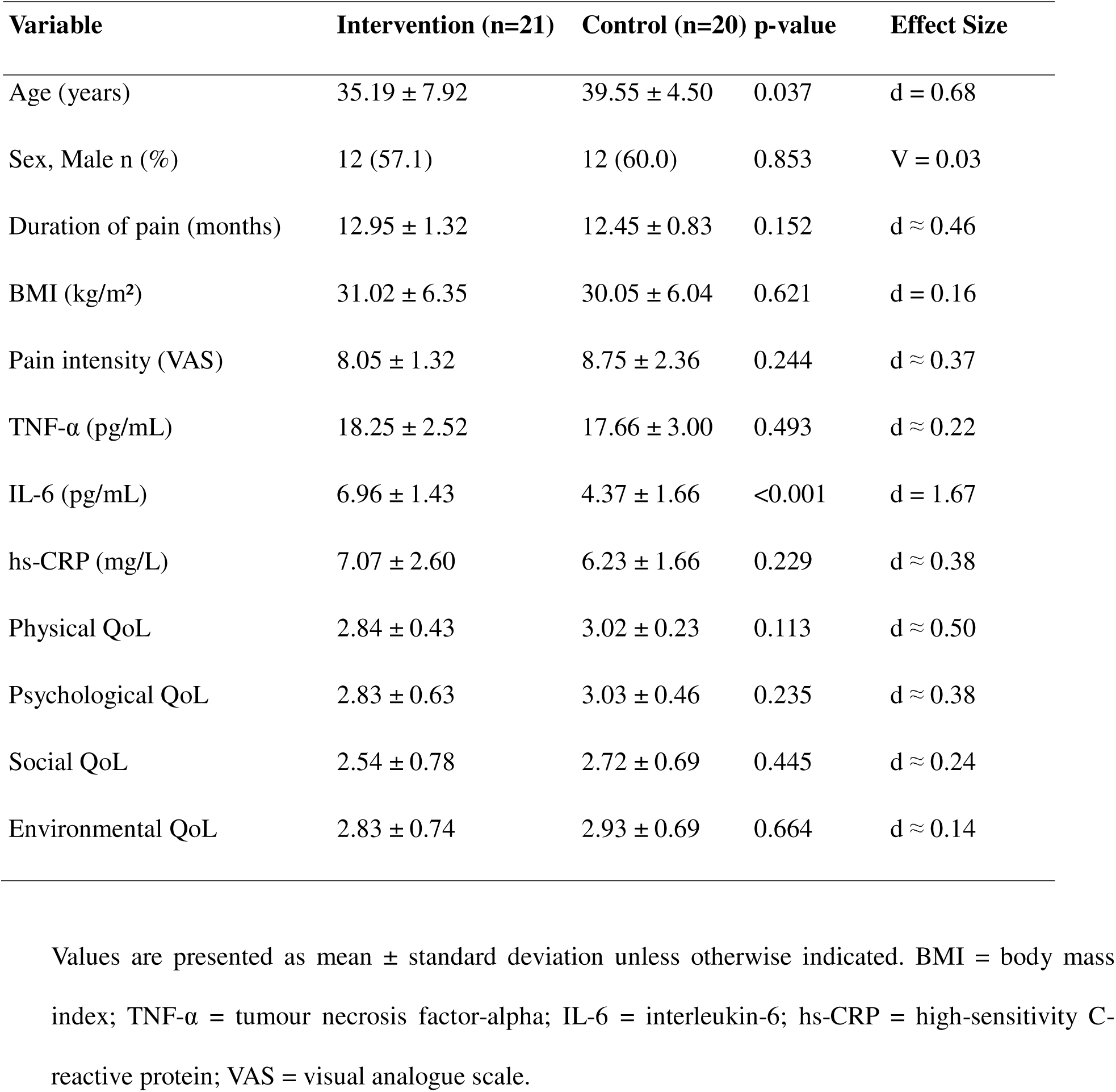
Baseline Demographic and Clinical Characteristics of Participants.

### Associations Between Changes in Inflammatory Biomarkers, Pain Intensity, and Quality of Life

Correlation analyses are presented in Table 2. Improvements in IL-6 (r = 0.434, p = 0.005) and hs-CRP (r = 0.444, p = 0.004) demonstrated moderate positive associations with improvements in pain intensity. No significant associations were observed between improvements in TNF-α and pain intensity (r = 0.236, p = 0.138). Similarly, changes in inflammatory biomarkers were not significantly associated with improvements in physical, psychological, social, or environmental quality-of-life domains (all p > 0.05).

**Table 2.** Associations Among Changes in Inflammatory Biomarkers, Pain Intensity, and Quality-of-Life Outcomes (N = 41)

| Variable | Imp_Pain | Imp_Physical | Imp_Psychological | Imp_Social | Imp_Environmental |
| --- | --- | --- | --- | --- | --- |
| Imp_TNF | 0.236 (p = 0.138) | -0.030 (p = 0.853) | 0.023 (p = 0.885) | -0.124 (p = 0.438) | 0.051 (p = 0.752) |
| Imp_IL6 | 0.434** (p = 0.005) | 0.018 (p = 0.910) | 0.035 (p = 0.829) | 0.081 (p = 0.614) | 0.091 (p = 0.573) |
| Imp_CRP | 0.444** (p = 0.004) | -0.297 (p = 0.059) | -0.287 (p = 0.069) | -0.241 (p = 0.130) | -0.222 (p = 0.162) |

Note. Imp = improvement score (Week 12 – Baseline); TNF-α = tumour necrosis factor-alpha; IL-6 = interleukin-6; hs-CRP = high-sensitivity C-reactive protein. Positive values indicate greater improvement. \*\*p < 0.01.

### Predictors of Improvement in Physical Quality of Life

The results of the multiple linear regression analysis predicting improvement in physical quality of life are presented in Table 3. Multiple linear regression analysis was conducted to determine whether changes in inflammatory biomarkers and pain intensity predicted improvement in the physical domain of quality of life after adjustment for age and treatment allocation. The overall model was statistically significant (F(6, 34) = 4.219, p = 0.003), explaining 42.7% of the variance in improvement in physical quality of life (R² = 0.427; adjusted R² = 0.326).

**Table 3.**
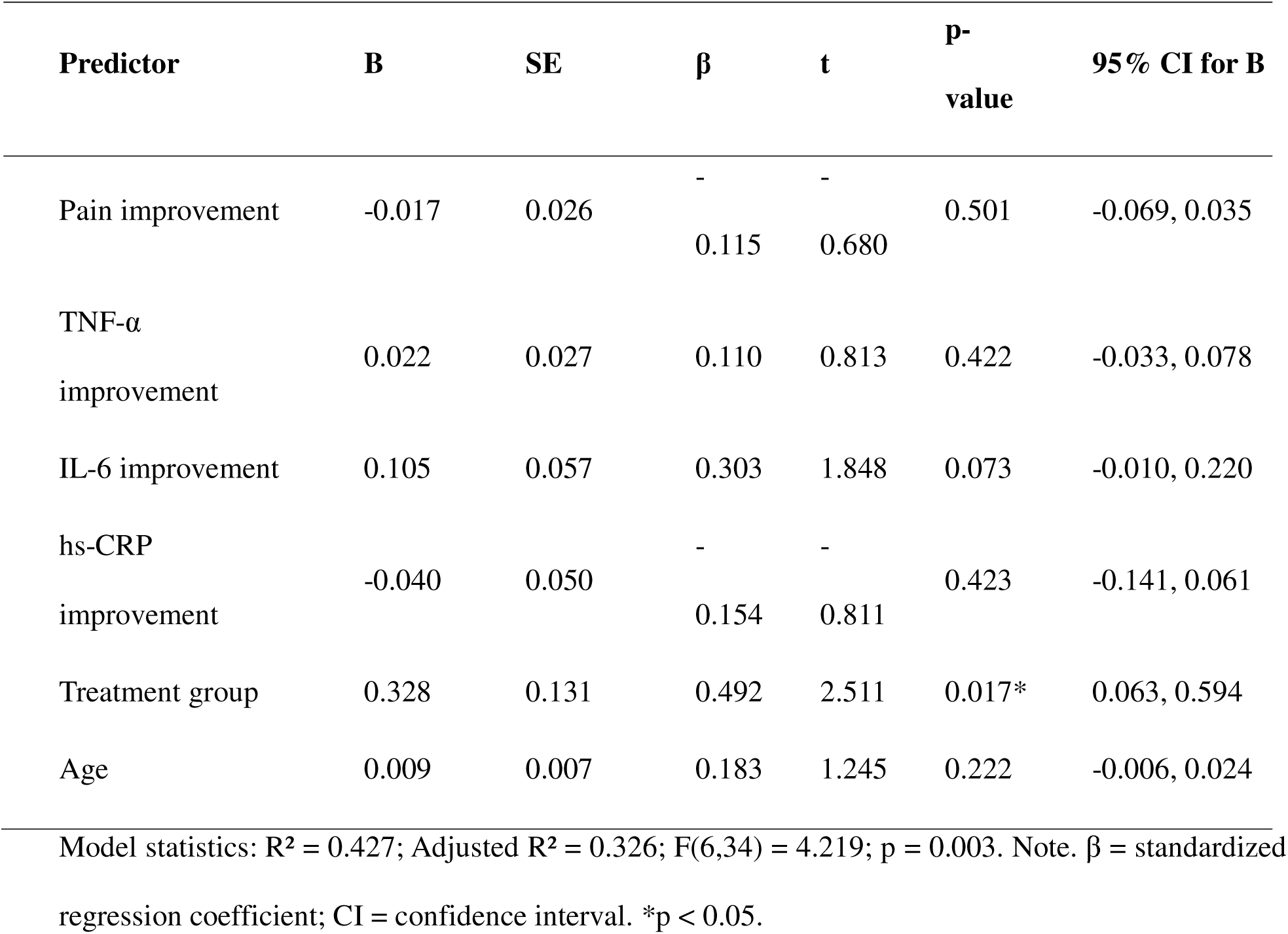
Multiple Linear Regression Analysis Predicting Improvement in Physical Quality of Life (N = 41)

| Predictor | B | SE | $\beta$ | t | p-value | 95% CI for B |
| --- | --- | --- | --- | --- | --- | --- |
| Pain improvement | -0.017 | 0.026 | -<br>0.115 | -<br>0.680 | 0.501 | -0.069, 0.035 |
| TNF-α<br>improvement | 0.022 | 0.027 | 0.110 | 0.813 | 0.422 | -0.033, 0.078 |
| IL-6 improvement | 0.105 | 0.057 | 0.303 | 1.848 | 0.073 | -0.010, 0.220 |
| hs-CRP<br>improvement | -0.040 | 0.050 | -<br>0.154 | -<br>0.811 | 0.423 | -0.141, 0.061 |
| Treatment group | 0.328 | 0.131 | 0.492 | 2.511 | 0.017* | 0.063, 0.594 |
| Age | 0.009 | 0.007 | 0.183 | 1.245 | 0.222 | -0.006, 0.024 |

Model statistics: $R^2 = 0.427$ ; Adjusted $R^2 = 0.326$ ; $F(6,34) = 4.219$ ; $p = 0.003$ . Note. $\beta$ = standardized regression coefficient; CI = confidence interval. \* $p < 0.05$ .

Treatment allocation emerged as the only significant independent predictor of improvement in physical quality of life (β = 0.492, p = 0.017). Improvements in IL-6 demonstrated a trend towards significance (β = 0.303, p = 0.073), whereas improvements in pain intensity (β = −0.115, p = 0.501), TNF-α (β = 0.110, p = 0.422), hs-CRP (β = −0.154, p = 0.423), and age (β = 0.183, p = 0.222) were not significant predictors. These findings suggest that participation in the aerobic exercise intervention was more strongly associated with improvement in physical quality of life than changes in inflammatory biomarkers or pain intensity (Table 3).

### Predictors of Improvement in Pain Intensity

The results of the multiple linear regression analysis predicting improvement in pain intensity are presented in Table 4. Multiple linear regression analysis was conducted to identify predictors of improvement in pain intensity following the intervention period. The overall model was statistically significant (F(5, 35) = 4.853, p = 0.002), explaining 40.9% of the variance in pain improvement (R² = 0.409; adjusted R² = 0.325).

**Table 4.** Multiple Linear Regression Analysis Predicting Improvement in Pain Intensity (N = 41)

| Predictor | B | SE | $\beta$ | t | p-value | 95% CI for B |
| --- | --- | --- | --- | --- | --- | --- |
| TNF-α improvement | 0.123 | 0.180 | 0.093 | 0.687 | 0.497 | -0.241, 0.488 |
| IL-6 improvement | 0.675 | 0.355 | 0.297 | 1.899 | 0.066 | -0.047, 1.396 |
| hs-CRP improvement | -0.064 | 0.327 | -0.037 | -0.194 | 0.847 | -0.727, 0.600 |
| Treatment group | -2.256 | 0.774 | -0.512 | -2.915 | 0.006* | -3.827, -0.685 |
| Age | 0.053 | 0.047 | 0.161 | 1.116 | 0.272 | -0.043, 0.149 |

Model statistics: $R^2 = 0.409$ ; Adjusted $R^2 = 0.325$ ; $F(5,35) = 4.853$ ; $p = 0.002$ . Note. $\beta$ = standardized regression coefficient; CI = confidence interval. \* $p < 0.05$ .

Treatment allocation was the only significant independent predictor of improvement in pain intensity (β = −0.512, p = 0.006). Improvements in IL-6 demonstrated a trend towards significance (β = 0.297, p = 0.066), whereas improvements in TNF-α (β = 0.093, p = 0.497), hs-CRP (β = −0.037, p = 0.847), and age (β = 0.161, p = 0.272) were not significant predictors. These findings indicate that assignment to the aerobic exercise intervention was the strongest determinant of pain improvement, while changes in inflammatory biomarkers showed limited independent contributions after adjustment for age and treatment allocation (Table 4).

No evidence of problematic multicollinearity was observed among the predictor variables included in the regression models, with variance inflation factor (VIF) values ranging from 1.09 to 2.14.

## Discussion

This secondary analysis examined the associations among changes in inflammatory biomarkers, pain intensity, and health-related quality of life (HRQoL) following a 12-week aerobic exercise programme in individuals with non-specific chronic low back pain (NSCLBP). Four principal findings emerged. First, improvements in interleukin-6 (IL-6) and high-sensitivity C-reactive protein (hs-CRP) were significantly associated with improvements in pain intensity in unadjusted analyses. Second, changes in inflammatory biomarkers were not significantly associated with improvements in HRQoL domains. Third, treatment allocation was the strongest independent predictor of both pain improvement and physical HRQoL improvement. Finally, although changes in IL-6 demonstrated trends towards independently predicting improvements in pain intensity and physical HRQoL, these associations were attenuated after adjustment for age and treatment allocation and did not reach statistical significance.

### Associations Between Inflammatory Biomarkers and Pain Improvement

The significant associations observed between improvements in IL-6, hs-CRP, and pain intensity support previous evidence suggesting that inflammatory processes are linked to pain experiences in individuals with NSCLBP. Elevated concentrations of pro-inflammatory biomarkers, particularly IL-6 and CRP, have been associated with greater pain severity, disability, and poorer clinical outcomes in chronic low back pain populations [4,5]. The present findings suggest that reductions in inflammatory activity were associated with concurrent improvements in pain intensity during rehabilitation.

Several biological mechanisms may explain these relationships. Pro-inflammatory cytokines contribute to peripheral nociceptor sensitisation, neuroimmune interactions, and alterations in central pain-processing pathways that may amplify pain perception and contribute to symptom persistence [4]. Consequently, lower inflammatory activity may be associated with reduced pain intensity and improved symptom experiences.

An important finding, however, was that the significant associations observed in the unadjusted analyses were attenuated after adjustment for age and treatment allocation. This suggests that intervention-related factors may account for a substantial proportion of the observed relationships. Therefore, while inflammatory biomarkers were associated with pain improvement, the present findings do not support inflammation as an independent predictor of pain improvement after accounting for these factors.

Improvements in TNF-α were not significantly associated with pain improvement. Although TNF-α has been implicated in chronic pain mechanisms, previous studies have reported less consistent associations between TNF-α concentrations and clinical outcomes than those observed for IL-6 and hs-CRP [5]. The absence of a significant association in the present study may reflect biological variability, limited statistical power, or the possibility that TNF-α is less responsive to physiological adaptations occurring during rehabilitation.

### Associations Between Inflammatory Biomarkers and Health-Related Quality of Life

Contrary to expectations, changes in inflammatory biomarkers were not significantly associated with improvements in any HRQoL domain. These findings suggest that reductions in inflammatory activity alone may not adequately explain broader improvements in patient-reported outcomes among individuals with NSCLBP.

HRQoL is a multidimensional construct influenced by physical functioning, emotional well-being, social participation, environmental factors, and personal perceptions of health. Improvements in these domains may therefore occur through mechanisms extending beyond biological changes in systemic inflammation. Previous research has highlighted the importance of psychological, behavioural, and social factors in determining HRQoL outcomes among individuals with chronic pain [6].

The absence of significant associations between biomarker changes and HRQoL outcomes may indicate that inflammation is only one component of a complex biopsychosocial process. Improvements in HRQoL may be more strongly influenced by factors such as increased physical activity, enhanced movement confidence, reduced fear-avoidance behaviour, improved self-efficacy, and greater participation in daily activities than by changes in inflammatory biomarkers alone [6,20,21]. Furthermore, the absence of statistically significant independent associations should be interpreted cautiously because the study may have been underpowered to detect small-to-moderate effects.

### Predictors of Pain and Physical Health-Related Quality-of-Life Improvement

Treatment allocation emerged as the strongest independent predictor of both pain improvement and physical HRQoL improvement. Participants assigned to the aerobic exercise group experienced greater improvements than those in the control group, even after adjustment for age and changes in inflammatory biomarkers.

This finding is consistent with previous evidence demonstrating the effectiveness of aerobic exercise for the management of chronic low back pain. Systematic reviews have reported that aerobic exercise can reduce pain intensity and improve physical functioning, disability, and quality of life through a combination of physiological, psychological, and behavioural mechanisms [20,21]. The present findings extend this literature by suggesting that the benefits associated with aerobic exercise may not be fully explained by reductions in systemic inflammation. Aerobic exercise may influence clinical outcomes through several complementary pathways, including improvements in cardiovascular fitness, physical conditioning, movement confidence, self-efficacy, and psychological well-being [20,21]. Exercise participation may also reduce fear-avoidance behaviours, encourage greater engagement in physical activity, and improve functional capacity, thereby contributing to improvements in pain and HRQoL [20,21]. These mechanisms are consistent with the biopsychosocial model of chronic pain, which emphasises the interaction of biological, psychological, and social influences on health outcomes [6].

Although improvements in IL-6 demonstrated trends towards predicting improvements in pain intensity and physical HRQoL, these relationships did not achieve statistical significance after adjustment for age and treatment allocation. Consequently, inflammatory changes may partially explain observed improvements in clinical outcomes, but they do not appear to be the primary determinants of treatment response in the present study.

### Clinical Implications

The findings have several implications for rehabilitation practice. The observed associations between improvements in IL-6, hs-CRP, and pain intensity suggest that inflammatory biomarkers may provide useful insight into biological changes accompanying symptom improvement in individuals with NSCLBP. However, the absence of significant relationships between inflammatory biomarkers and HRQoL indicates that clinicians should not rely solely on biological markers when evaluating treatment effectiveness.

Instead, rehabilitation programmes should continue to target multiple dimensions of health, including pain reduction, physical functioning, psychological well-being, self-efficacy, and social participation. The findings further support the inclusion of aerobic exercise as a valuable component of multidisciplinary rehabilitation programmes for individuals with chronic low back pain.

### Strengths and Limitations

This study has several strengths. It utilised data derived from a randomized controlled trial, incorporated both objective inflammatory biomarkers and patient-reported outcomes, achieved full participant retention throughout the intervention period, and examined multiple domains of HRQoL. The use of regression analyses enabled exploration of the independent contributions of biomarker changes and treatment allocation to clinical outcomes.

Several limitations should be acknowledged. First, the sample size was determined for the parent randomized controlled trial rather than for the correlation and regression analyses performed in this secondary study, potentially limiting statistical power to detect smaller associations. Second, significant baseline differences were observed for age and IL-6 concentrations despite randomization. Although age and treatment allocation were included in regression models, residual confounding cannot be excluded. Furthermore, the observed baseline imbalance in IL-6 concentrations may have influenced the magnitude of change observed during the intervention period and should be considered when interpreting the findings.

Potential confounding by unmeasured factors, including medication use, body mass index, physical activity levels outside the intervention, smoking status, and comorbidities, cannot be excluded. These factors may influence inflammatory biomarker concentrations and clinical outcomes and therefore should be considered when interpreting the findings.

Third, the analyses were based on change scores, which may be susceptible to measurement error and regression-to-the-mean effects. Furthermore, change-score analyses do not fully account for baseline differences and may not adequately capture the longitudinal relationships among inflammatory biomarkers, pain intensity, and HRQoL. Fourth, the exploratory nature of the analyses and the multiple statistical comparisons performed increase the possibility of Type I error. Fifth, treatment allocation may have captured intervention-related influences beyond exercise exposure alone, including therapist interaction, participant engagement, adherence, and treatment expectations. The parent trial was retrospectively registered because of administrative delays. Although the protocol and outcome measures were established before participant recruitment, retrospective registration may influence perceptions of methodological transparency. Finally, participants were recruited from a single tertiary healthcare institution, potentially limiting the generalisability of the findings.

### Future Research

Future studies using larger samples, prospective longitudinal designs, and repeated biomarker assessments are needed to clarify the temporal relationships among systemic inflammation, pain intensity, and HRQoL in individuals with NSCLBP. Future investigations should also incorporate repeated-measures analytical approaches rather than relying solely on change scores and should evaluate potential psychological and behavioural factors, including self-efficacy, fear-avoidance beliefs, physical activity levels, and treatment expectations. Adequately powered studies may also explore whether inflammatory biomarkers contribute directly or indirectly to improvements in patient-centred outcomes following rehabilitation interventions.

### Conclusion

Among individuals with NSCLBP who participated in a 12-week intervention programme, improvements in IL-6 and hs-CRP were significantly associated with improvements in pain intensity, whereas changes in inflammatory biomarkers were not significantly associated with improvements in HRQoL outcomes. Treatment allocation emerged as the strongest predictor of both pain improvement and physical HRQoL improvement. These exploratory findings suggest that changes in systemic inflammation may be associated with improvements in pain intensity but do not appear to fully explain broader improvements in HRQoL. Further research is required to clarify the complex relationships among inflammation, pain, and patient-centred outcomes in chronic low back pain.

## Data Availability

All data produced in the present study are available upon reasonable request to the authors

## DECLARATIONS

### Ethics approval and consent to participate

Ethical approval was obtained from the Rivers State University Teaching Hospital Health Research Ethics Committee (Approval No.: RSUTH/REC/2025897). Written informed consent was obtained from all participants prior to enrolment. The study was conducted in accordance with the ethical principles outlined in the Declaration of Helsinki.

### Consent for publication

Not applicable. This study does not contain any individual person’s data in any form (including individual details, images, or videos).

### Availability of data and materials

The de-identified participant dataset supporting the findings of this study, together with the statistical analysis code and data dictionary, are available from the corresponding author upon reasonable request and subject to institutional ethical approval requirements.

### Clinical trial registration

Pan African Clinical Trials Registry (PACTR), Registration Number: PACTR202606887164888; retrospectively registered on 11 June 2026. The study protocol, eligibility criteria, intervention procedures, and outcome measures were established prior to participant recruitment.

### Protocol availability

The study protocol and statistical analysis plan are available from the corresponding author upon reasonable request.

### Patient and public involvement

Patients and members of the public were not involved in the design, conduct, reporting, or dissemination plans of this research.

### Authors’ contributions

- Conceptualization: VN, CE
- Methodology: VN, CE, CO, OA, KE
- Investigation (Data Collection): VN, AM, OA, KE
- Formal Analysis: VN, OA, KE
- Data Interpretation: VN, OA, KE
- Writing – Original Draft: VN, OA, KE and AN
- Writing – Review & Editing: VN, CO, CE and AN
- Supervision: CE
- Project Administration: VN
- Guarantor: VN
- Approval of Final Manuscript: All authors reviewed and approved the final manuscript and agree to be accountable for all aspects of the work.

## Acknowledgements

The authors sincerely acknowledge all participants who volunteered to take part in this study. Appreciation is extended to the physiotherapists, laboratory personnel, and research assistants whose contributions facilitated participant recruitment, intervention delivery, and data collection.

## List of Abbreviations

BMI: Body Mass Index
CONSORT: Consolidated Standards of Reporting Trials
ELISA: Enzyme-Linked Immunosorbent Assay
HRQoL: Health-Related Quality of Life
HRR: Heart Rate Reserve
hs-CRP: High-Sensitivity C-Reactive Protein
IL-6: Interleukin-6
NSCLBP: Non-Specific Chronic Low Back Pain
PACTR: Pan African Clinical Trials Registry
RCT: Randomized Controlled Trial
RPE: Rating of Perceived Exertion
SD: Standard Deviation
TNF-α: Tumour Necrosis Factor-Alpha
VAS: Visual Analogue Scale
WHOQOL-BREF: World Health Organization Quality of Life Brief Questionnaire

